# Show us the Data: Global COVID-19 Wastewater Monitoring Efforts, Equity, and Gaps

**DOI:** 10.1101/2021.03.14.21253564

**Authors:** Colleen C. Naughton, Fernando A. Roman, Ana Grace F. Alvarado, Arianna Q. Tariqi, Matthew A. Deeming, Kyle Bibby, Aaron Bivins, Joan B. Rose, Gertjan Medema, Warish Ahmed, Panagis Katsivelis, Vajra Allan, Ryan Sinclair, Yihan Zhang, Maureen N. Kinyua

## Abstract

A year since the declaration of the global coronavirus disease 2019 (COVID-19) pandemic there were over 110 million cases and 2.5 million deaths. Learning from methods to track community spread of other viruses such as poliovirus, environmental virologists and those in the wastewater based epidemiology (WBE) field quickly adapted their existing methods to detect SARS-CoV-2 RNA in wastewater. Unlike COVID-19 case and mortality data, there was not a global dashboard to track wastewater monitoring of SARS-CoV-2 RNA worldwide. This study provides a one year review of the “COVIDPoops19” global dashboard of universities, sites, and countries monitoring SARS-CoV-2 RNA in wastewater. Methods to assemble the dashboard combined standard literature review, direct submissions, and daily, social media keyword searches. Over 200 universities, 1,000 sites, and 55 countries with 59 dashboards monitor wastewater for SARS-CoV-2 RNA. However, monitoring is primarily in high-income countries (65%) with less access to this valuable tool in low and middle income countries (35%). Data are not widely shared publicly or accessible to researchers to further inform public health actions, perform meta-analysis, better coordinate, and determine equitable distribution of monitoring sites. For WBE to be used to its full potential during COVID-19 and beyond, show us the data.

## 1. Introduction

In one year, the coronavirus disease 2019 (COVID-19) pandemic has resulted in 110 million cases and 2.5 million deaths worldwide (Dong et al., 2020). When the novel coronavirus strain (SARS-CoV-2) that causes COVID-19 emerged in late 2019, environmental virologists began rapidly adapting their methods from those supporting surveys of other pathogens within wastewater (GWPP, 2021) including use of public health elements to address concerns associated with monitoring SARS-CoV-2 RNA in wastewater. Some of the first major monitoring efforts for SARS-CoV-2 in wastewater were in the Netherlands (Lodder and de Roda Husman, 2020; Medema et al., 2020b), Australia (Ahmed et al., 2020), Italy (La Rosa et al., 2020), and the United States (Sherchan et al., 2020). A global coordination effort was proposed to share and standardize sampling strategies, virus recovery methodologies, and data for WBE for SARS-CoV-2 (Bivins et al., 2020). COVID-19 Wastewater Based Epidemiology (WBE) and environmental/wastewater surveillance or monitoring are being used to describe this effort and has grown from just a few countries in March 2020 to at least 55 countries and over 200 universities a year later (Naughton et al., 2021).

Both the growth and recognition of WBE for SARS-CoV-2 monitoring has been rapid and widespread. Wastewater monitoring to address epidemiological questions has been used historically at mostly smaller scales to track enteric viruses and other pathogens (GWPP, 2021) including the poliovirus vaccine and wildtype strains (Hovi et al., 2011), norovirus, adenovirus, and other pathogens (Ali et al., 2021), antimicrobial resistance (Hendriksen et al., 2019), and drugs such as opioids (Burgard et al., 2014; Li et al., 2019; Schmidt, 2020). Because of the COVID-19 pandemic, a year later, at least seven countries, Finland (THL, 2021), France (Obépine, 2021), Hungary (NNK), Luxembourg (LIST, 2021), Netherlands (Rijksoverheid, 2021), Spain (VATar, 2021), and Turkey (Kocamemi et al., 2020), have nationalized wastewater monitoring for SARS-CoV-2. The United States (NWSS, 2021) and Canada (CWN, 2021) have established national coordination networks/systems. At least four countries have regional level monitoring: Australia (Victoria, 2021; Queensland, 2021), Brazil (ANA, 2021), South Africa (SAMRC, 2021), Switzerland (EWAG, 2021a, EWAG, 2021b), and the United Kingdom (SEPA, 2021). Throughout these countries and globally, newspaper, online and television outlets have extensively covered SARS-CoV-2 wastewater monitoring with local to national politicians calling for widespread application of wastewater testing. Whereas COVID-19 case and death data has been widely available globally, such as through the Johns Hopkins University dashboard (Dong et al., 2020), even the locations of COVID-19 wastewater testing are less available and difficult to track.

Though challenges exist to standardize wastewater testing methods and data normalization (Medema et al., 2021a), public health departments (CWN, 2020), utilities, scientists and engineers have an ethical obligation, especially during a pandemic, to provide this information to the public who is being monitored. The goal of this study is to provide a global dashboard and year one analysis of SARS-CoV-2 wastewater testing to inform the public (general population, public health departments, municipalities, and researchers) where this type of testing is taking place and provide links to available data for decision making and better coordination. Our hypothesis was that much of the wastewater SARS-CoV-2 data will not be publicly available and low and middle income countries would have less access to wastewater monitoring. This study uses the “COVIDPoops19” dashboard to identify gaps in wastewater monitoring to make recommendations for science communication of wastewater data, and as a call to action for more forthcoming and transparent open data sharing.

## 2. Materials and Methods

To create a global dashboard of reported wastewater monitoring efforts, six different data sources were used (see Supporting Information Figure S1): (1) the COVID-19 WBE website (COVID-19 WBE Collaborative, 2021), (2) webinars, (3) Google form submissions, (4) literature searches, (5) Twitter keyword searches, and (6) Google keyword searches. ArcGIS Online Dashboards was chosen as the host platform (ESRI, 2020). First, points were added from the COVID-19 WBE collaborative publication map as country points (COVID-19 WBE Collaborative, 2021). A link to a Google form was made available at the bottom of the COVIDPoops19 dashboard for users to submit public data points. A Twitter account (@COVIDPoops19) was created for the dashboard and the UC Merced co-authors performed key word searches daily for six combinations of ‘wastewater’ or ‘sewage’ and ‘COVID19’ or ‘COVID-19’ or ‘SARS-CoV-2’.

From advertisements on Twitter and the United States National Science Foundation (NSF) COVID-19 WBE Research Coordination Network (RCN) (Wastewater Surveillance RCN, 2021), the co-authors regularly attended webinars to learn about different monitoring efforts. Only publicly reported locations and data from websites and news articles were added to the dashboard. Google was used to check for missing U.S. states and territories. For example, a combination of “Puerto Rico” and “wastewater”, “sewage”, “monitoring” and “COVID-19” and “SARS-CoV-2” keywords were used to see whether there were missing articles that were not found by the daily keyword searches on Twitter.

Wastewater monitoring locations for SARS-CoV-2, news articles, publications, Google form submissions, dashboard/data and other web links were collected and sorted into four categories: (1) dashboard/data, (2) university, (3) country, and (4) sites. GPS coordinates in WGS 84 coordinate system for the dashboard were either directly extracted when provided or approximated from the location mentioned in the source. If a city, county, or country were found testing their wastewater for SARS-CoV-2 without specific sampling sites mentioned, then a point was placed near the centroid of the mentioned area tested to associate the testing site with a location. When other public dashboards for wastewater testing efforts provided coordinates for their sampled sites, those were downloaded and utilized as site points on the COVIDPoops19 dashboard. The COVIDPoops19 dashboard was usually updated weekly depending on the number of points gathered and submitted.

Although keyword and literature searches were predominantly in English, the dashboard team includes English, French, and Spanish speakers, and the dashboard had a broad submission from international stakeholders via the Google form as well as engagement during international webinars. Many researchers in other countries also publish and post in English.

After the collection of sites, universities, and countries, the spatial distribution of wastewater monitoring was analyzed. Countries were sorted based on the World Bank income classifications (high income, upper middle income, lower middle income, and low income) (World Bank, 2021). ArcGIS Pro 2.6.1 was used to map the number of sites and universities monitoring wastewater for SARS-CoV-2 globally. With a large number of sites and universities monitoring SARS-CoV-2 in wastewater, the United States was chosen to further classify based on the fifty states and five inhabited territories. The U.S. Centers for Disease Control and Prevention (CDC) rankings based on COVID-19 cases, hospitalization, and deaths were compared to entities without wastewater monitoring for SARS-CoV-2 (CDC, 2021b).

Dashboards were categorized based on their presentation, communication style, and data availability. Results of SARS-CoV-2 testing in wastewater were presented as maps, graphs, a small written description or solely by color (demonstrating an increase or decrease of trend). Dashboard communication style categories were: video, FAQ page, a short written format (less than three paragraphs), longer descriptions (three or more paragraphs), and no form of written communication. The simplicity of the communication was also determined by whether the description given was: (1) technical, more specifics on the science behind SARS-CoV-2 wastewater testing (included information on lab processes), or (2) a simpler form of communication that would be understandable to the general public (used general vocabulary to inform as to why wastewater is being employed to test for SARS-CoV-2). Dashboards were checked for whether they provided downloadable data, the file type, and the variables available.

## 3. Results and Discussion

As of March 11th, 2021, a year after declaration of the COVID-19 pandemic (Cucinotta and Vanelli, 2020), the COVIDPoops19 global dashboard for wastewater monitoring of SARS-CoV-2 included 235 universities, 59 dashboards, and 1,488 sites in 55 countries. Between September 2020 and March 11th, 2021, there were 60 submissions on the Google form linked to the COVIDPoops19 dashboard. Since the dashboard was published publicly in September 2021, there have been 25,679 visits. The COVIDPoops19 twitter account has acquired over 2,000 followers between May 2020 and March 2021.

Of the 195 countries in the world (U.S. DOS, 2021), 55 contain wastewater monitoring. Of these 55, 36 (65%) are in high-income countries, 11 (20%) are upper middle income, 8 (15%) are lower middle income, and 0% are low income countries (Figure 1). Similar to COVID-19 individual testing and Personal Protective Equipment (PPE) (Kavanagh e al., 2020, McMahon et al., 2020) and vaccination efforts (Lancet Commission, 2021), access to wastewater testing is also more widely available in high income countries.

**Figure 1:**
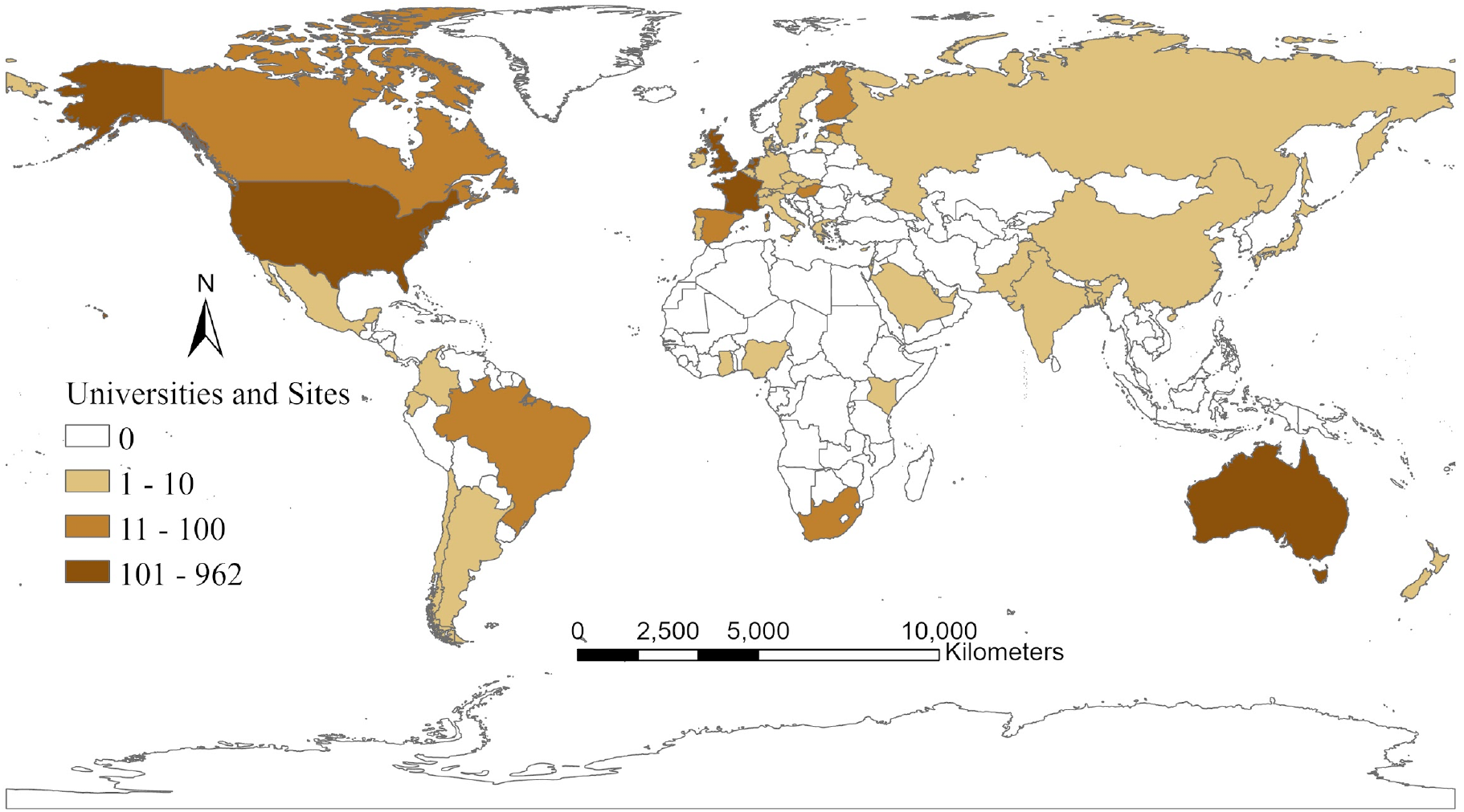
World map with countries using wastewater monitoring of SARS-CoV-2.

The COVIDPoops19 dashboard is extensive but is likely an underestimate of the locations testing wastewater for SARS-CoV-2 RNA because it is limited to publicly available data. Many private companies who are monitoring wastewater for SARS-CoV-2 are limited by what their client(s) (e.g., public health department, municipality, etc.) allow to be shared. For example, Biobot Analytics is a private company that conducts WBE (Biobot Analytics, 2021). Biobot has processed wastewater from at least 300 sites in 42 states in the United States (Wiggins, 2020). Some Biobot sites were found and posted from news articles and publicly available dashboards, e.g., Eastern Massachusetts (Biobot Analytics, 2021b), Chattanooga (Biobot Analytics, 2021c), Nantucket (Town & Country of Nantucket, 2021), Delaware (Biobot Analytics, 2021d), etc., but the COVIDPoops19 dashboard was missing other sites. In May 2021, Biobot did release aggregate data (Biobot Analytics, 2021e), over a year into monitoring. Similarly, universities do not publicly report all the sites they are sampling from.

The United States (U.S.) had the highest number of universities and sites (962) monitoring for SARS-CoV-2 globally. Of the 50 States and five inhabited territories of the U.S. and the District of Columbia, there was no record of wastewater testing for SARS-CoV-2 RNA in: (1) American Samoa, (2) Guam, (3) Puerto Rico, (4) U.S. Virgin Islands, and (5) Northern Mariana Islands (see Figure 2).

**Figure 2:**
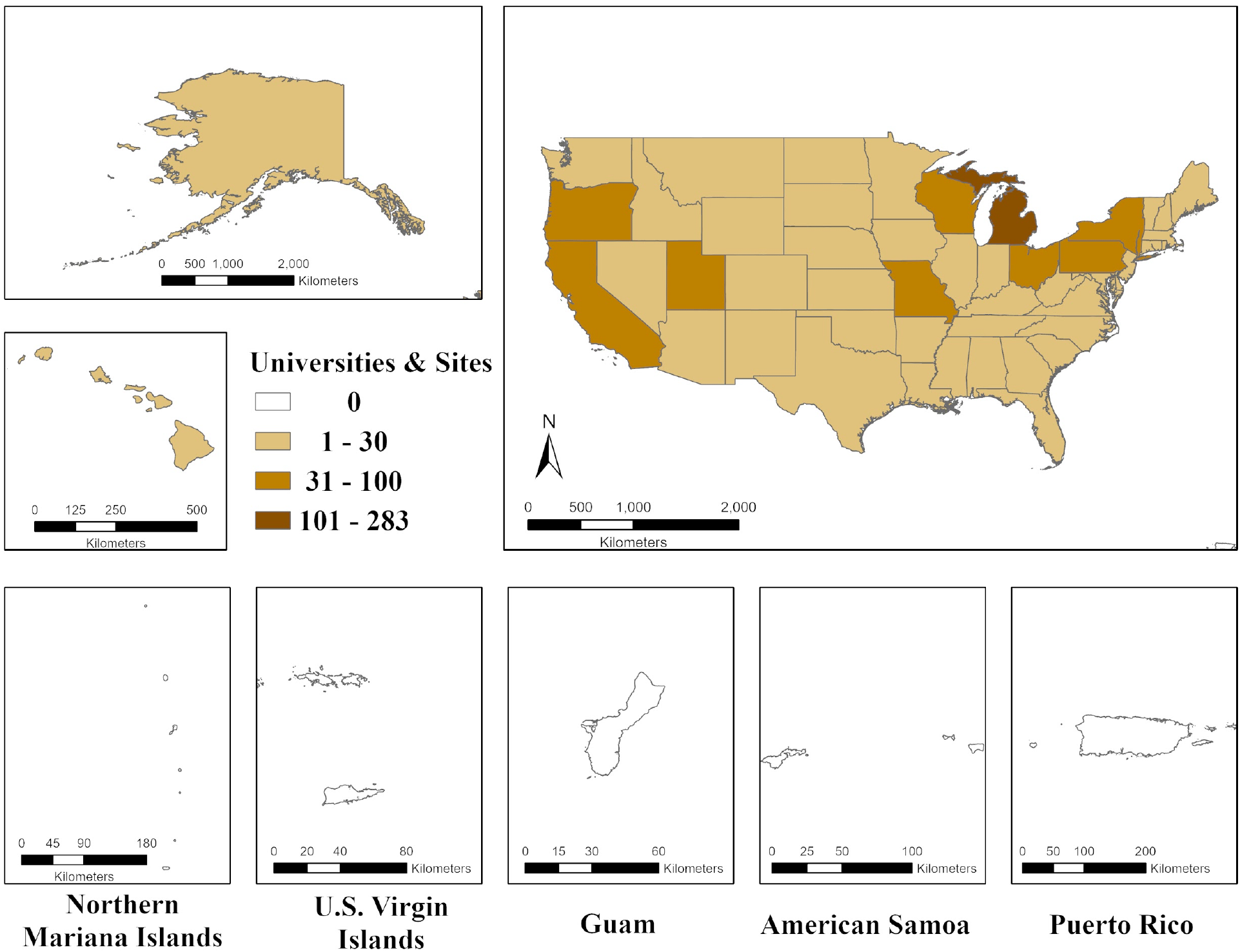
Map of the United States and inhabited territories testing wastewater for SARS-CoV-2 RNA.

While the lack of wastewater monitoring may not be directly related to the number of cases and deaths within the state, WBE has potential as an early warning system and to identify hotspots to better target public health measures to prevent further COVID-19 cases (Ahmed et al., 2021). Iowa had no publicly disclosed wastewater testing until the University of Iowa added testing in February 2021 (University of Iowa, 2021). Iowa ranked eighth in cases per 100,000 people and sixteenth in deaths per 100,000 people in the U.S. (CDC, 2021b). South Dakota had only one location monitoring for SARS-CoV-2 RNA since July 2020 and is ranked second highest for total cases per 100,000 people and seventh highest for deaths per 100,000 people within the U.S. as of February 15th, 2021 (CDC, 2021b). There was no record of wastewater testing for SARS-CoV-2 RNA in any of the inhabited territories within the U.S. However, the U.S. territories all ranked in the bottom 10 among U.S. states and territories with the least number of cases and deaths per 100,000 people based on available testing (CDC, 2021b).

Despite over 200 universities, 1,000 sites, and 55 countries with reported wastewater surveillance for SARS-CoV-2, there are a limited number of entities that make their data openly accessible with only 59 publicly available dashboards. Of these 59 dashboards, only 18 had downloadable data for further analysis (see Supporting Information Table S1). Data are downloadable as .csv, .xlsx, .rda, .pdf, or .pitemx files depending on the dashboard. Typical data include flow rates, collection dates, coordinates, days since sampled, sample types, gene copy information, and if the virus was observed in the sample. Data available and units vary for each dashboard, as there is no common data standard followed among the different endeavors.

Fifty-nine dashboards were categorized on how their results were primarily presented. Twenty-eight (47%) presented their dashboards in the form of a map, 28 (47%) used graphs, two (3%) solely gave a written description of the results (Erie County, 2021; Lewis and Clark County, 2021), and one (2%) presented an image with a color to demonstrate the trend (Indiana Borough, 2021). Fourteen (24%) dashboards used both a graph and a map. Eighteen dashboards (30%) used colors to visually present results.

Fourteen of the 59 dashboards (34%) had no description of the data provided. Of the 45 dashboards that had some form of description, 25 (56%) dashboards used a short written format, 13 (29%) included more than three written paragraphs, nine (20%) included a Frequently Asked Questions (FAQ) page, and three (7%) had videos. Five dashboards (13%) used a combination of communication styles. Valencia, Spain had a video and included multiple paragraphs (GoAigua, 2020). New Haven, Connecticut and Bozeman, Montana both had a video and a short written format to describe SARS-CoV-2 testing in wastewater (Yale University, 2020; Healthy Gallatin, 2021), and the Luxembourg and Missouri dashboards have a short written format and a section with FAQ (LIST, 2021; Missouri Department of Health and Senior Services, 2021). Lastly, of the dashboards that presented some form of communication, 34 (76%) were written in language that could be understood by the general public, whereas 11 (24%) had very specific and detailed scientific information. For example, the dashboard used for Valencia, Spain had a simple communication style for a general audience to understand how wastewater can be used as a tool to better understand COVID-19 trends in their area (GoAigua, 2020). In contrast, the dashboard used for Minas Gerais, Brazil went more in-depth with the scientific specifics of the lab results and was categorized as more technical (ANA, 2021).

While offering detailed and technical information about the wastewater testing process/protocol is ideal, it is also important to communicate the benefits of wastewater testing for the general public. For this reason, successful communication styles should include more understandable vocabulary (e.g., less scientific jargon) with links to WBE case studies, while offering links to more detailed information for more technical audiences (e.g., researchers, public health departments, and municipalities). Additionally, providing a video explanation can help more visual learners.

It is essential to ensure appropriate public health surveillance systems and open data access in pandemic response. The Canadian Water Network states, “During a public health emergency, it is imperative that all parties involved in surveillance share data in a timely fashion.” (CWN, 2020) Providing open access to data collected from testing wastewater for SARS-CoV-2 RNA, along with effective communication and properly handling sensitive information, can better inform the public which will allow for a collective fight against the COVID-19 pandemic.

Increased access to wastewater testing data could provide other researchers, such as data scientists, the opportunity to further develop algorithms, compare between sites, and better analyze the data to make it more useful to inform public health decisions instead of keeping it internal. Individuals may use wastewater data in their personal risk decisions if they see an increase in concentrations in the wastewater in their area or where they may travel. However, an ongoing challenge of WBE is the lack of normalization across datasets. This is a nascent research space with high variability in methods used to collect, process, and analyze samples.

Increased data sharing may allow for analysis across collection sites and to identify which methods work best in high income countries (HIC) and lower middle income countries (LMIC) settings (Pandey et al., 2021). Wastewater testing could be a useful and cost effective option in low-resource settings with limited clinical testing (Hart and Halden, 2020; Usman et al., 2020). Greater open data would also facilitate better collaboration, coordination, and equity analysis. Most testing is concentrated in HICs. However, even within HICs there may be inequity in distribution to high-income, urban areas with less diversity, similar to disparities in individual testing (Hopper et al., 2020) and vaccination (KFF, 2021) in the U.S.

The U.S. National Wastewater Surveillance System (NWSS) currently only allows access to the wastewater data on their internal dashboard to public health departments (CDC, 2021a). The United States Health and Human Services (HHS) recently aimed to test 30% of the U.S. population through wastewater (Genomeweb, 2020). HHS has yet to publicly release the locations where wastewater sampling occurred. Without knowing all the locations, researchers, the media, and the general public have no way to determine if wastewater testing is equitably distributed among the 50 states, territories, and low-income, minority, and rural communities.

The United States has an OPEN (Open, Public, Electronic, and Necessary) Government Data Act that mandates federal agencies to make their data open (Data.gov, 20201). Fifty-three other countries that also have open data websites and policies are listed on Data.gov. The European Council prioritized the adoption of Open Science and reusability of research data, promoting FAIR (Findable, Accessible, Interoperable, and Reusable) data principles (Mons et al., 2017). COVID-19 case and death data has been invaluable during the pandemic to inform the public and policies. Wastewater data can be aggregated and de-identified similar to case, hospitalization, and death data to protect private health information.

For WBE to be used to its full potential as a public health tool during and after the COVID-19 pandemic, data must be more openly shared with the public and among researchers. Wastewater monitoring and support for dashboard development must also be expanded to lower income countries and areas. Wastewater monitoring will remain important throughout vaccination efforts to monitor for outbreaks (Smith et al., 2020) and can be used to track the spread of variants at larger scales (Martin et al., 2021) only if they show us the data.

## Data Availability

All data is available publicly through ArcGIS online. A link to our public dashboard is here: https://arcg.is/1aummW

## Supporting Information

Figure of COVIDPoops19 dashboard workflow and table of wastewater dashboards for COVID-19 and whether they included downloadable data.

## Acknowledgements

We would like to thank all those contributing to the global effort to monitor SARS-CoV-2 RNA in wastewater that made the COVIDPoops19 dashboard and this publication possible. Funding sources include: a Center for Information Technology Research in the Interest of Society (CITRIS) COVID-19 emergency seed grant, Bill and Melinda Gates Foundation, and National Science Foundation (NSF) grants # 2037834, #2027752, and #2038087. Any opinions, findings, and conclusions or recommendations expressed in this material are those of the author(s) and do not necessarily reflect the views of the National Science Foundation or other funding agencies.

## References

Agência Nacional de Águas (ANA). (2021). “MONITORAMENTO COVID ESGOTOS.” Secretaria de Estado de Saúde de Minas Gerais. https://coronavirus.saude.mg.gov.br/transparencia/monitoramento-covid-esgotos

Ahmed, W., Angel, N., Edson, J., Bibby, K., Bivins, A., O’Brien, J. W., Choi, P. M., Kitajima, M., Simpson, S. L., Li, J., Tscharke, B., Verhagen, R., Smith, W. J. M., Zaugg, J., Dierens, L., Hugenholtz, P., Thomas, K. V., Mueller, J. F. First confirmed detection of SARS-CoV-2 in untreated wastewater in Australia: a proof of concept for the wastewater surveillance of COVID-19 in the community. Science of the Total Environment. 2020, 728: 138764.

Ahmed, W., Tscharke, B., Bertsch, P.M., Bibby, K., Bivins, A., Choi, P., Clarke, L., Dwyer, J., Edson, J., Nguyen, T.M.H, O’Brien, J.W., Simpson, S.L., Sherman, P., Thomas, K.V., Verhagen, R., Zaugg, J., Mueller, J.F. SARS-CoV-2 RNA monitoring in wastewater as a potential early warning system for COVID-19 transmission in the community: A temporal case study. Science of the Total Environment. 2021, 761: 144216.

Ali, W., Zhang, H., Wang, Z., Chang, C., Javed, A., Ali, K., Du, W., Niazi, N.Z., Mao, K., Yang, Z. Occurrence of various viruses and recent evidence of SARS-CoV-2 in wastewater systems. Journal of Hazardous Materials. 2021, 414 (15): 125439. DOI: 10.1016/j.jhazmat.2021.125439

Biobot Analytics. (2021a). https://www.biobot.io/about_usBiobot

Analytics. (2021b). “Wastewater COVID-19 Tracking.” Massachusetts Water Resources Authority. https://www.mwra.com/biobot/biobotdata.htm

Biobot Analytics. (2021c). “Biobot Analytics Report.” The Hamilton County Health Department. https://connect.chattanooga.gov/covid-biobot-analysis-reports/

Biobot Analytics. (2021d). “New Castle County Confirmed COVID-19 Cases (Rolling 7-Day Average).” https://compassred.shinyapps.io/ncco_wastewater/

Biobot Analytics. (2021e). “Nationwide Wastewater Monitoring Network.” https://biobot.io/data/

Bivins, A., North, D., Ahmad, A., Ahmed, W., Alm, E., Been, F., et al. Wastewater-Based Epidemiology: Global Collaborative to Maximize Contributions in the Fight Against COVID-19. Environmental Science and Technology. 2020, DOI: 10.1021/acs.est.0c02388

Burgard, D.A., Banta-Green, C., Field, J.A. Working Upstream: How Far Can You Go with Sewage-Based Drug Epidemiology? Environmental Science and Technology. 2014, 48 (3): 1362–1368.

Canadian Water Network (CWN). (y2020). “Ethics and communications guidance for wastewater surveillance to inform public health decision-making about COVID-19.” https://cwn-rce.ca/wp-content/uploads/COVID19-Wastewater-Coalition-Ethics-and-Communications-Guidance-v4-Sept-2020.pdf

Canadian Water Network (CWN). (2021). COVID-19 Wastewater Coalition. https://cwn-rce.ca/covid-19-wastewater-coalition/

Centers for Disease Control and Prevention (CDC). (2021a). “National Wastewater Surveillance System (NWSS).” Centers for Disease Control and Prevention. https://www.cdc.gov/coronavirus/2019-ncov/cases-updates/wastewater-surveillance.html

Centers for Disease Control and Prevention (CDC). (2021b). “United States COVID-19 Cases and Deaths by State.” Centers for Disease Control and Prevention. https://covid.cdc.gov/covid-data-tracker/#cases_casesper100klast7days

COVID-19 WBE Collaborative. (2021). “COVID-19 WBE Collaborative.” www.COVID19wbec.org

Cucinotta, D. and Vanelli, M. WHO Declares COVID-19 a Pandemic. Acta Biomed. 2020, 91(1): 157–160, DOI: 10.23750/abm.v91i1.9397

Data.gov. (2021). “Open Government.” https://www.data.gov/open-gov/

Dong, E., Duh, H., Gardner, L. An interactive web-based dashboard to track COVID-19 in real time. Lancet Inf Dis. 2020, 20 (5): 533–534, DOI: 10.1016/S1473-3099(20)30120-1

Erie County. (2021). Tag: Biobot. Erie County Pennsylvania. https://eriecountypa.gov/tag/biobot/

ESRI Inc. (2020). ArcGIS Online (Version 2.6). Esri Inc. www.esri.com/en-us/arcgis/products/arcgis-online/overview

Gallatin City-County Health Department (Healthy Gallatin). (2021). “Wastewater Testing and COVID-19.” Healthy Gallatin. https://www.healthygallatin.org/coronavirus-covid-19/wastewater-data/

Genomeweb. (2020). “AquaVitas Partners with HHS, CDC on Wastewater Coronavirus Study.” Genomeweb. https://www.genomeweb.com/infectious-disease/aquavitas-partners-hhs-cdc-wastewater-coronavirus-study#.YD4ElpNKibs

Global Water Pathogens Project. (y2021). https://www.waterpathogens.org/

GoAigua. (2020). “Valencia anticipates coronavirus outbreaks thanks to GoAigua SARS Analytics.” Idrica. https://www.idrica.com/blog/valencia-anticipates-coronavirus-outbreaks-thanks-to-goaigua-sars-analytics/

Hart, O.E., Halden, R.U. Computational analysis of SARS-Cov-2/COVID-19 Surveillance by wastewater-based epidemiology locally and globally: Feasibility, economy, opportunities and challenges. Science of the Total Environment. 2020, 730: 138875, DOI: 10.1016/j.scitotenv.2020.138875.

Hendriksen, R.S., Munk, P., Njage, P., van Bunnik McNally, L. et al. Global monitoring of antimicrobial resistance based on metagenomics analyses of urban sewage. Nature Communications. 2019, 10 (1124): 1124, DOI: 10.103/s41467-019-08853-3.

Hopper, M.W., Napoles, A.M., Perz-Stable, E.J. COVID-19 and Racial/Ethnic Disparities. JAMA. 2020, 323 (24): 2466–2467. DOI: 10.1001/jama.2020.8598.

Hovi, T., Shulman, L.M., Van der Avoort, H., Deshpande, J., Roivainen, M., De Gourville, E.M. Role of environmental poliovirus surveillance in polio eradication and beyond. Epidemiology & Infection. 2011, 140 (1): 1–13, DOI: 10.1017/S095026881000316X

Kaiser Family Foundation (KFF). (2021). “Early State Vaccination Data Raise Warning Flags for Racial Equity.” KFF Coronavirus (COVID-19) Policy Watch. January 21, 2021. Authors: Ndugga, N, Pham, O., Hill, L., Artiga, S., Megistu, S. https://www.kff.org/policy-watch/early-state-vaccination-data-raise-warning-flags-racial-equity/

Kavanagh, M., Erondu, N.A., Tomori, O., Dzau, V.J., Okiro, E. Maleche, A. Aniebo, I.C., Rugege, U., Holms, C.B., Gostin, L.O. Access to Life-Saving Medical Resources for African Countries: COVID-19 Testing and Response, Ethics, and Politics. The Lancet. 2020, 395 (10238): 1735–1738, DOI: 10.1016/S0140-6736(20)31093-X

Kocamemi, B. A., Kurt, H., Sait, A., Kadi, H., Sarac, F., Aydin, I., Saatci, A. M., Pakdemirli, B. Nationwide SARS-CoV-2 Surveillance Study for Sewage and Sludges of Wastewater Treatment Plants in Turkey. medRxiv. 2020, DOI: 10.1101/2020.11.29.20240549

Indiana Borough. (2021). Weekly Trend Report Wastewater Surveillance. Indiana County, PA. https://www.indianaboro.com/news/categories/wastewater-surveillance

La Rosa, G., Iaconelli, M., Mancini, P., Ferraro, G.B., Veneri, C., Bonadonna, L., Lucentini, L., Suffredini, E. First detection of SARS-CoV-2 in untreated wastewaters in Italy. Science of the Total Environment. 2020, 736 (20): 139652, DOI: 10.1016/j.scitotenv.2020.139652

Lancet Commission on COVID-19 Vaccines and Therapeutics Task Force Members (Lancet Commission). Urgent needs of low-income and middle-income countries for COVID-19 vaccines and therapeutics. The Lancet. 2021, 397 (10274): 562–564. DOI: 10.1016/S0140-6736(21)00242-7

Li, X., Du, P., Zhang, W., Zhang, L. Wastewater: A new resource for the war against illicit drugs. Current Opinion in Environmental Science & Health. 2019, 9: 73–76, DOI: 10.1016/j.coesh.2019.05.003

Lodder, W., de Roda Husman, A.M. SARS-CoV-2 in wastewater: potential health risk, but also data source. Lancet Gastroenterol Hepatol. 2020, 5 (6): 533–534, DOI: 10.1016/S2468-1253(20)30087-X

Luxembourg Institute of Science and Technology (LIST). (2021). “MONITORING THE EVOLUTION OF COVID-19 IN WASTEWATER.” Luxembourg Institute of Science and Technology. https://www.list.lu/en/covid-19/coronastep/

Martin, J., Klapsa, D., Wilton, T., Zambon, M., Bentley, E., Bujaki, E., Fritzsche, M., Mate, R., Majumdar, M. Tracking SARS-CoV-2 in Sewage: Evidence of Changes in Virus Variant Predominance during COVID-19 Pandemic. Viruses. 2021, 12: 1144. DOI: 10.3390/v12101144

McMahon, D.E., Peters, G.A., Ivers, L.C., Freeman, E.E. Global resource shortages during COVID-19: Bad news for low-income countries. PLoS Negl Trop Dis. 2020, 14 (7): e0008412, DOI: 10.1371/journal.pntd.0008412

Medema, G., Been, F., Heijnen, L., Petterson, S. Implementation of environmental surveillance for SARS-CoV-2 virus to support public health decisions: Opportunities and Challenges. Curr Opin Environ Sci Health. 2020a, 17: 49–71, DOI: 10.1016/j.coesh.2020.09.006

Medema, G., Heijnen, L., Elsinga, G., Italiaander, R., Brouwer, A. Presence of SARS-Coronavirus-2 RNA in Sewage and Correlation with Reported COVID-19 Prevalence in the Early Stage of the Epidemic in The Netherlands. Environ. Sci Technol. Letters. 2020b, 7: 511–516, DOI: 10.1021/acs.estlett.0c00357

Missouri Department of Health and Senior Services. (2021). “The Sewershed Surveillance Project.” Missouri Department of Health and Senior Services. https://storymaps.arcgis.com/stories/f7f5492486114da6b5d6fdc07f81aacf

Mons, B., Cameron, N., Velterrop, J., Dumontier, M., da Silva Santos, L.O.B, Wilkinson, M.D. ‘Cloudy, increasingly FAIR; revisiting the FAIR Data guiding principles for the European Open Science Cloud. Information Services & Use. 2017, 37(1): 49–56, DOI: 103233/ISU-170824.

Naughton, C.C., Roman, F.A.R., Tariqi, A.Q., Kadonsky, K.K. (2021). COVIDPoops19 Summary of Global SARS-CoV-2 Wastewater Monitoring Efforts by UC Merced Researchers. ArcGIS Online Dashboard. Created September 2021. https://arcg.is/1aummW

Nemzeti Népegészségügyi Központ (NNK). (2021). “COVID-19 Tájékoztató Oldal.” https://www.nnk.gov.hu/index.php/koronavirus-tajekoztato/743-jelentos-emelkedes-a-megbetegedesek-szamaban-a-kovetkezo-1-2-hetben-tovabbra-sem-varhato

Observatoire ÉPIdémiologique dans les Eaux usées (Obépine). (2021). “Rapport d’analyses: présentation des résultats.” https://www.reseau-obepine.fr/donnees-ouvertes/

Pandey, D., Verma, S., Verma, P., Mahanty, B., Dutta, K., Daverey, A., Arunachalam, K. SARS-CoV-2 in wastewater: Challenges for Developing Countries. International Journal of Hygiene and Environmental Health. 2021, 231: 113634, DOI: 10.1016/j.ijheh.2020.113634.

Queensland Government. (2021). “Wastewater surveillance program results.” https://www.qld.gov.au/health/conditions/health-alerts/coronavirus-covid-19/current-status/wastewater

Rijksoverheid. (2021). “Virusdeeltjes in rioolwater.” https://coronadashboard.rijksoverheid.nl/landelijk/rioolwater

SAMRC. (2021). SAMRC SARS-CoV-2 Wastewater Surveillance Dashboard: An Early Warning System for COVID-19 Infections. https://www.samrc.ac.za/wbe/

Schmidt, C. Watcher in the Wastewater. Nat Biotechnol. 2020, 38: 917–920, DOI: 10.1038/s41587-020-0620-2

Scottish Environment Protection Agency (SEPA). (2021). “Current National Overview.” https://informatics.sepa.org.uk/RNAmonitoring/

Sherchan, S.P, Shahin, S., Ward, L.M., Tandukar, S., Aw, T.G., Schmitz, B., Ahmed, W., Kitajima, M. First Detection of SARS-CoV-2 RNA in wastewater in North America: A Study in Lewis and Clark County. (2021). Local COVID-19 Decision-Making Dashboard. Lewis and Clark County Montana. https://www.lccountymt.gov/health/covid-19/decision-making-dashboard.html

Sherchan, S.P, Shahin, S., Ward, L.M., Tandukar, S., Aw, T.G., Schmitz, B., Ahmed, W., Kitajima, M. First Detection of SARS-CoV-2 RNA in wastewater in North America: A Study in Louisiana, USA. Science of the Total Environment. 2020, 743: 140621, DOI: 10.1016/j.scitotenv.2020.140621

Smith, T., Cassell, G., Bhatnagar, A. Wastewater Surveillance Can Have a Second Act in COVID-19 Vaccine Distribution. JAMA Health Forum. 2020, DOI 10.1001/jamahealthforum.2020.1616.

Swiss Federal Institute of Aquatic Science and Technology (EAWAG). (2021a). “WWTP Zürich Werdhölzli -raw influent.” http://parsivel-eawag.ch/sarscov2/ARA_Werdhoelzli_ddPCR.html

Swiss Federal Institute of Aquatic Science and Technology (EAWAG). (2021b). “WWTP Lausanne-Vidy - raw influent.” https://sensors-eawag.ch/sarscov2/STEP_Vidy_ddPCR.html

Terveyden ja hyvinvoinnin laitos (THL). (2021). “Koronaviruksen jätevesiseurannan viikkoraportti.” https://www.thl.fi/episeuranta/jatevesi/jatevesiseuranta_viikkoraportti.html

Town & Country of Nantucket, MA. (2021) “Surfside Wastewater Treatment Facility Covid-19 Results.” https://nantucket-ma.gov/1864/Surfside-Wastewater-Treatment-Facility-C

University of Iowa. (2021). “Campus update: Vaccine update; virtual spring commencement; wastewater testing pilot; variant confirmed in Johnson County; campus operations; self-reported testing data.” https://coronavirus.uiowa.edu/news/2021/02/campus-update-vaccine-update-virtual-spring-commencement-wastewater-testing-pilot

U.S. Department of State (DOS)/Bureau of Intelligence and Research. (2021). “Independent States in the World.” U.S. Department of State. https://www.state.gov/independent-states-in-the-world/

Usman, M., Farooq, M., Hanna, K. Existence of SARS-CoV-2 in Wastewater: Implications for Its Environmental Transmission in Developing Countries. Environmental Science and Technology. 2020, 54 (13): 7758–7759. DOI: 10.1021/acs.est.0c02777.

VATar COVID-19. (2021). Cuadro de mando de evolucion de EDAR. Ministerio para la Transicion Ecologica y el Reto Demografico. https://miteco.maps.arcgis.com/apps/opsdashboard/index.html#/aab0e0653d694289b310f6485f9f2226

Victoria State Government. (2021). “Wastewater testing.” https://www.dhhs.vic.gov.au/wastewater-testing-covid-19

Wastewater Surveillance for SARS-CoV-2 NSF Research Coordination Network (RCN). (2021). https://sites.nd.edu/rcn-wastewater-sarscov2/

Wiggins, O. (2020). “Flushing out the coronavirus: Universities, cities and states are testing wastewater for the virus.” The Washington Post. December 25, 2020. https://www.washingtonpost.com/local/md-politics/wastewater-coronavirus-testing-/2020/12/25/f83330ce-4011-11eb-8db8-395dedaaa036_story.html

World Bank. (2021). “World Bank Country and Lending Groups.” The World Bank Data. https://datahelpdesk.worldbank.org/knowledgebase/articles/906519-world-bank-country-and-lending-groups

Yale University. (2020). “Yale covid-19 Wastewater Tracker.” https://yalecovidwastewater.com/

