## Supplemental Information for "Show us the Data: Global COVID-19 Wastewater Monitoring Efforts, Equity, and Gaps"

**Table of Contents**

**Figure S1:** COVIDPoops19 dashboard data workflow …………..S2

**Table S1:** List of dashboards/data for wastewater monitoring of SARS-CoV-2 …………..S3

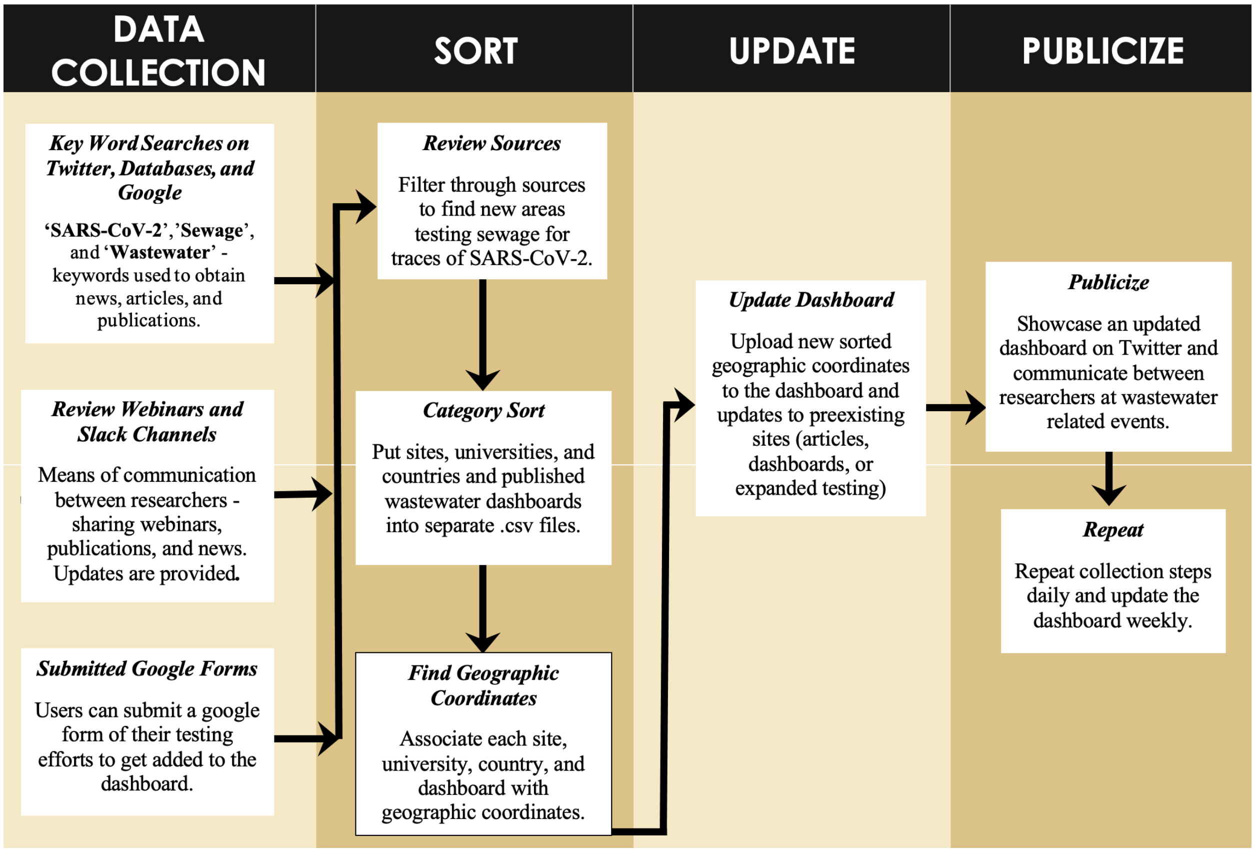


**Figure S1:** COVIDPoops19 dashboard data workflow

**Supporting Information Table S1:** List of dashboards/data for wastewater monitoring of SARS-CoV-2

| N | Country | City | State | Latitude | Longitude | Website | Downloadable Data? |
|---|---|---|---|---|---|---|---|
| 1 | United States | Athens | Georgia | 33.9519 | -83.3576 | https://lipplab-uga.github.io/covid_wastewater_lipplab_athens/ | Yes |
| 2 | United States | New Haven | Connecticut | 41.3083 | -72.9279 | https://yalecovidwastewater.com/ | No |
| 3 | United States | Clemson | South Carolina | 34.6834 | -82.8374 | https://www.clemson.edu/covid-19/testing/wastewater-dashboard.html | No |
| 4 | United States | | Utah | 39.321 | -111.09 | https://deq.utah.gov/water-quality/sars-cov-2-sewage-monitoring | No |
| 5 | United States | Tempe | Arizona | 33.4255 | -111.94 | https://covid19.tempe.gov/ | Yes |
| 6 | United States | | Massachusetts | 42.3506 | -70.9577 | http://www.mwra.com/biobot/biobotdata.htm | Yes |
| 7 | United States | | Ohio | 40.4173 | -82.9071 | https://coronavirus.ohio.gov/wps/portal/gov/covid-19/dashboards/wastewater | Yes |
| 8 | United States | New Castle | Delaware | 39.6955 | -75.6734 | http://biobot.newcastlede.gov/ | No |
| 9 | United States | Boise | Idaho | 43.615 | -116.2023 | https://www.cityofboise.org/departments/mayor/coronavirus-covid-19-information/covid-19-data/wastewater-testing/ | Yes |
| 10 | United States | Bozeman | Montana | 45.677 | -111.0429 | https://www.healthygallatin.org/coronavirus-covid-19/wastewater-data/ | No |
| 11 | Netherlands | | | 52.1326 | 5.2913 | https://coronadashboard.rijksoverheid.nl/ | Yes |
| 12 | Spain | Valencia | | 39.4699 | -0.3763 | https://www.idrica.com/blog/valencia-anticipates-coronavirus-outbreaks-thanks-to-goaigua-sars-analytics/ | No |
| 13 | Canada | Ottawa | Ontario | 45.4215 | -75.6972 | https://613covid.ca/wastewater/# | Yes |
| 14 | Hungary | | | 47.1683 | 19.5032 | https://www.nnk.gov.hu/index.php/koronavirus-tajekoztato/743-jelentos-emelkedes-a-megbetegedesek-szamaban-a-kovetkezo-1-2-hetben-tovabbra-sem-varhato | No |
| 15 | Finland | | | 61.9241 | 25.7482 | https://www.thl.fi/episeuranta/jatevesi/jatevesiseuranta_viikkoraportti.html | Yes |

| N | Country | City | State | Latitude | Longitude | Website | Downloadable Data? |
|---|---|---|---|---|---|---|---|
| 16 | United States | Gilbert | Arizona | 33.3528 | -111.789 | https://experience.arcgis.com/experience/19a447509ff44fcf922ad0892b7b2cce/page/page_21/ | No |
| 17 | United Kingdom | | Scotland | 56.8169 | -4.1827 | https://informatics.sepa.org.uk/RNAmonitoring/ | Yes |
| 18 | Switzerland | Zurich | | 47.3961 | 8.545548 | http://parsivel-eawag.ch/sarscov2/ARA_Werdhoelzli_ddPCR.html | Yes |
| 19 | United States | Corvallis | Oregon | 44.5646 | -123.262 | http://people.oregonstate.edu/~levit/Software/CorvallisWastewater102120.html | No |
| 20 | Australia | | Victoria | -37.809 | 144.970 | https://www.dhhs.vic.gov.au/wastewater-monitoring-covid-19 | No |
| 21 | Brazil | | Rio Grande do Sul | -29.186 | -51.170 | https://app.powerbi.com/view?r=eyJrIjoiMzNhZjNiNjUtZjQxZS00ZDA0LWFhZTctNjNlNjg0Mjg5Y2FiIiwidCI6IjE1ZGNkOTA5LThkYzAtNDBlOS1hMWU1LWNlY2IwNTNjZGQxYSJ9&pageName=ReportSection | No |
| 22 | South Africa | | | -33.924 | 18.4232 | https://www.samrc.ac.za/wbe/ | No |
| 23 | Luxembourg | | | 49.6101 | 6.130619 | https://www.list.lu/en/covid-19/coronastep/ | No |
| 24 | United States | Indiana | Pennsylvania | 40.6231 | -79.155 | https://www.indianaboro.com/news/categories/wastewater-surveillance | No |
| 25 | Spain | | Catalonia (region) | 41.983 | 2.82493 | https://sarsaigua.icra.cat/ | Yes |
| 26 | United States | Nantucket (Island) | Massachusetts | 41.2667 | -70.0158 | https://nantucket-ma.gov/1864/Surfside-Wastewater-Treatment-Facility-C | No |
| 27 | United States | Keene | New Hampshire | 42.9263 | -72.2832 | https://www.keene.edu/featured/fall2020/covid-19-dashboard/ | No |
| 28 | United States | Burlington | Vermont | 44.4880 | -73.2080 | https://coronavirus-response-burlingtonvt.hub.arcgis.com/pages/wastewater-monitoring | No |

| N | Country | City | State | Latitude | Longitude | Website | Downloadable Data? |
|---|---|---|---|---|---|---|---|
| 29 | United States | | Oregon | 44.0578 | -120.7331 | https://public.tableau.com/profile/oregon.health.authority.covid.19#!/vizhome/OregonsSARS-CoV-2WastewaterMonitoring/WastewaterDashboard | No |
| 30 | United States | Jupiter | Florida | 26.9245 | -80.10546 | https://loxahatcheeriver.org/wastewater-surveillance/ | No |
| 31 | Australia | | Queensland | -21.631 | 148.03938 | https://www.qld.gov.au/health/conditions/health-alerts/coronavirus-covid-19/current-status/wastewater | No |
| 32 | United States | | Wisconsin | 43.7844 | -88.7879 | https://www.dhs.wisconsin.gov/covid-19/wastewater.htm | Yes |
| 33 | Brazil | | Minas Gerais | -19.912 | -44.0282 | https://t.co/RY4D877qeW?amp=1 | No |
| 34 | United States | Erie | Pennsylvania | 42.1292 | -80.0851 | https://eriecountypa.gov/tag/biobot/ | No |
| 35 | United States | | Michigan | 44.7648 | -84.7052 | https://egle.maps.arcgis.com/apps/webappviewer/index.html?id=8e3cb66aca204876a1db0e1b663af805 | Yes |
| 36 | United States | Chattanooga | Tennessee | 35.0528 | -85.3330 | https://connect.chattanooga.gov/covid-biobot-analysis-reports/ | No |
| 37 | United States | | Pennsylvania | 41.2892 | -77.44583 | https://www.cor.pa.gov/Pages/COVID-19.aspx | No |
| 38 | United States | Southeastern | Virginia | 37.0299 | -76.3452 | https://www.hrsd.com/HRSD-COVID-19-Surveillance | No |
| 39 | United States | | Colorado | 39.7392 | -104.9903 | https://cdphe.maps.arcgis.com/apps/opsdashboard/index.html#/d79cf93c3938470ca4bcc4823328946b | Yes |
| 40 | United States | | Illinois | 41.6441 | -88.450 | https://portal.rjngroup.com/arcgisportal/apps/opsdashboard/index.html#/594d4b1b2dd840958cedb50b1381982b | No |
| 41 | United States | | Missouri | 38.5762 | -92.175 | https://storymaps.arcgis.com/stories/f7f5492486114da6b5d6fdc07f81aacf | No |
| 42 | Switzerland | Lausanne | Vaud | 46.5244 | 6.59186 | https://sensors-eawag.ch/sarscov2/STEP_Vidy_ddPCR.html | Yes |
| 43 | United States | Cambridge | Massachusetts | 42.3742 | -71.1165 | https://cityofcambridge.shinyapps.io/COVID19/#shiny-tab-wastewater | No |

| N | Country | City | State | Latitude | Longitude | Website | Downloadable Data? |
|---|---|---|---|---|---|---|---|
| 44 | United States | | New Mexico | 32.631 | -105.86 | https://www.env.nm.gov/wastewater-surveillance-system-data-dashboard/ | Yes |
| 45 | Canada | Windsor | Ontario | 42.296 | -83.0255 | https://wechu.org/cv/weekly-epidemiological-summary | No |
| 46 | United States | San Diego | California | 32.8797 | -117.229 | https://returntolearn.ucsd.edu/dashboard/index.html | No |
| 47 | United States | Helena | Montana | 46.590 | -112.019 | https://www.lccountymt.gov/health/covid-19/local-covid-19-decision-making-dashboard.html | No |
| 48 | USA | Mansfield | Connecticut | 41.8114 | -72.264 | https://covid-testing.uconn.edu/dashboard/ | No |
| 49 | France | | | 46.6013 75 | 2.3095 | https://www.reseau-obepine.fr/donnees-ouvertes/ | No |
| 50 | Canada | Calgary | Alberta | 50.9940 | -114.071 | https://www.chi-csm.ca/ | No |
| 51 | Spain | Madrid | | 40.4200 | -3.7019 | https://www.canaldeisabelsegunda.es/mapa-vigia | No |
| 52 | Canada | | Northwest Territories | 64.8255 | -124.84 | https://nwt-covid.shinyapps.io/Testing-and-Cases/?lang=1 | No |
| 53 | Sweden | Uppsala | | 59.8586 | 17.69389 | https://crush-covid.shinyapps.io/crush_covid/ | No |
| 54 | United States | Telluride | Colorado | 37.9375 | -107.81 | https://sanmiguelco.maps.arcgis.com/apps/opsdashboard/index.html#/56e682135d1d4128bee1a0426aed1d10 | Yes |
| 55 | Canada | | Metro Vancouver | 49.2501 | -123.08 | http://www.metrovancouver.org/services/liquid-waste/environmental-management/covid-19-wastewater/Pages/default.aspx | No |
| 56 | United States | Honolulu | Hawaii | 21.3069 | -157.85 | https://www.oneoahu.org/dashboard | Yes |
| 57 | Spain | | | 40.4637 | -3.7492 | https://miteco.maps.arcgis.com/apps/opsdashboard/index.html#/aab0e0653d694289b310f6485f9f2226 | No |
| 58 | USA | St. Mary's County | Maryland | 38.3441 | -76.5764 | http://smchd.org/covid-19-wastewater | No |
| 59 | Spain | Edar Bens | A Coruna | 43.3677 | -8.4559 | Edarbens.es/covid19 | No |
